# Speech therapy combined with Cerebrolysin in enhancing aphasia recovery after acute ischemic stroke: ESCAS pilot study

**DOI:** 10.1101/2024.02.23.24303302

**Authors:** Volker Homberg, Dragoș Cătălin Jianu, Adina Stan, Ștefan Strilciuc, Vlad-Florin Chelaru, Michał Karliński, Michael Brainin, Wolf Dieter Heiss, Dafin Mureșanu, Pamela M Enderby

**Author notes:** **Correspondence:** Dafin F. Muresanu, No. 8 Victor Babes Street.

## Abstract

**Background:** Stroke-induced aphasia significantly impacts communication and quality of life. Despite the standard treatment being speech and language therapy (SLT), outcomes vary, highlighting the need for additional therapies. Cerebrolysin, a neuroprotective and neurotrophic agent, has shown potential in stroke management. This study examines the effectiveness of combining Cerebrolysin with SLT in treating post-stroke aphasia.

**Methods:** The ESCAS trial, a prospective, randomized-controlled, double-blinded phase 4 study, was conducted in two Romanian stroke centers. Participants included those with left middle cerebral artery territory ischemic stroke and Broca or mixed non-fluent aphasia, enrolled 3-5 days post-stroke. Inclusion criteria were right-handedness and Romanian as mother tongue; exclusion criteria were prior strokes, severe comprehension deficits, contraindications to MRI, and pre-existing neurodegenerative or psychiatric diseases. Participants received Cerebrolysin or a placebo combined with SLT in ten-day cycles over three intervals.

**Results:** Out of 132 enrolled patients, 123 were included in the Intention To Treat analysis, and 120 in the Per Protocol analysis. The Cerebrolysin group showed significant improvement in Western Aphasia Battery scores (p < 0.001) and National Institutes of Health Stroke Scale scores (p < 0.001). Modified Rankin Scale and Barthel index scores also improved, with notable differences at the final study visit (Day 90). Safety analysis raised no concerns.

**Conclusions:** Cerebrolysin combined with SLT offers promising potential for enhancing recovery in post-stroke aphasia. Significant improvements were observed in language and neurological deficits, underscoring the importance of adjunctive therapies in aphasia rehabilitation. Further research with larger cohorts is needed to fully establish the efficacy of this combination therapy.

**Clinical Trial Registration:** https://www.isrctn.com/ISRCTN54581790

## Introduction

Aphasia stands out as one of the most debilitating after ischemic stroke.^1,2^ It not only affects the individual’s ability to communicate but also has profound implications on their quality of life, self-esteem, and social interactions. Post-stroke aphasia can manifest in various forms, from difficulty in retrieving words to a complete loss of speech, understanding, reading, or writing.^3–5^

Speech and language therapy (SLT) remains the gold standard for aphasia rehabilitation. While many patients experience some degree of spontaneous recovery in the initial weeks to months following a stroke, the benefits of SLT in enhancing this recovery and ensuring long-term improvements are well-documented.^6^ However, the degree of recovery is variable, and about one third of stroke survivors continue to live with chronic aphasia.^7^ This variability underscores the need for adjunctive treatments that can potentiate the effects of SLT and offer hope to those with persistent communication deficits.^3^ Cerebrolysin, a biological agent containing a mixture of low-molecular-weight peptides, and amino acids, has been explored for its potential neuroprotective and neurotrophic effects due to its ability to mimic endogenous neurotrophic factors, thereby supporting both neuroprotection and neurorecovery.^8^ Beneficial effects of Cerebrolysin are attributed to its ability to reduce oxidative stress, inflammation, and apoptosis in the brain.^9^ Several clinical trials have investigated the therapeutic potential of Cerebrolysin in patients with neurological conditions, including acute ischemic stroke.^10–15^ These studies have reported improvements in clinical outcomes, reduced infarct volume, and enhanced neurological recovery. Cerebrolysin has been generally very well-tolerated in clinical trials and has been included in several clinical practice guidelines.^16–18^

The ESCAS trial was conceived to bridge this knowledge gap. By evaluating the synergistic effects of Cerebrolysin and SLT, the study aimed to provide insights that could improve the therapeutic approach to aphasia rehabilitation.

## Methodology

The ESCAS study was an exploratory, prospective, randomized-controlled, double-blinded, phase 4 academic study designed to assess the efficacy and safety of Cerebrolysin in combination with speech therapy compared to a placebo (saline solution) in combination with speech therapy for the treatment of aphasia following acute ischemic stroke. No concomitant use of Cerebrolysin for motor recovery was allowed. The trial was conducted between June 2020 to October 2022 in two Romanian stroke centers (Cluj County Emergency Hospital in Cluj-Napoca, and Pius Brînzeu County Emergency Hospital in Timisoara), enrolling consecutive eligible stroke patients, and was approved by the Ethics Committee of the Iuliu Hatieganu University of Medicine and Pharmacy (8 Babeş Street, 400012 Cluj-Napoca, Romania; ref: 122/24.03.2020), as well as local hospital IRB boards. The study, as well as the accompanying clinical study protocol and statistical analysis protocol, were prospectively registered in ISRCTN with the identifier ISRCTN54581790 (https://doi.org/10.1186/ISRCTN54581790).^19^

### Study inclusion criteria

- Radiological (either CT or MRI) and clinical confirmation of acute ischemic stroke in the left MCA territory.
- Presence of Broca or mixed non-fluent aphasia.
- Enrollment in the study between 3 and 5 days post-stroke.
- Right-handedness.
- Daily use of Romanian as the primary language.
- Provision of signed informed consent.

### Study exclusion criteria

- History of symptomatic ischemic or hemorrhagic stroke.
- Severe comprehension deficits that could compromise the understanding of informed consent or instructions, such as fluent aphasias (e.g., Wernicke aphasia) or global aphasias.
- History of epilepsy or EEG-documented epileptic discharges.
- Severe chronic renal or liver failure, indicated by AST, ALT levels greater than 4 times the normal values, or creatinine levels exceeding 4mg/dl.
- Presence of life-threatening diseases.
- Uncorrectable auditory or visual deficits that could impair testing.

### Study procedures

Upon initiation of the study, participants underwent their first visit, scheduled between 3 to 5 days post-stroke. This baseline visit (Day 0) involved the collection of participants’ demographics and medical history. Comprehensive assessments were conducted using tools such as the Western Aphasia Battery (WAB) and the National Institutes of Health Stroke Scale (NIHSS). A month into the study, specifically on Day 30 ± 3, participants returned for their second visit. During this session, evaluations were carried out using the WAB, NIHSS, Barthel Index (BI), and the modified Rankin Scale (mRS). Any adverse events (AE) or severe adverse events (SAE) experienced by the participants were also meticulously documented. Two months post-baseline, on Day 60 ± 3, the third visit took place. Participants underwent similar evaluations as the previous visit, including monitoring for any adverse or severe adverse events. The fourth and last visit was on Day 90 ± 3. This final session mirrored the assessments and monitoring procedures of the second and third visits. Parallel to the study visits, participants received a structured treatment regimen. They received an intravenous infusion of 30 ml Cerebrolysin (diluted in saline, total volume 250ml) or saline (250ml) and underwent one hour of speech therapy per day for ten days within two-week intervals, amounting to a total of three treatment cycles during the periods of study days 1-14, 29-42, and 57-70. The total amount of SLT for both study groups was 30 hours. The therapy sessions were conducted by neuropsychologists experienced in speech and language therapy, as no additional national level certification exists in this specific field. To ensure consistency in therapeutic approach and materials, the content and resources for speech therapy were standardized across all study centers using principles and practical applications described in the Aphasia Handbook by Ardila (2014). Throughout individual speech therapy sessions, patients received various worksheets to follow the entire recovery protocol and to cover a wide range of examples, enabling them to continue their recovery independently or with family support after the formal therapy sessions concluded. Intensity of the regimen was adapted to suit patient needs and content was personalized to account for other barriers such as literacy, based on the best judgement of the therapist. Given the constraints imposed by the pandemic, some sessions were conducted remotely to minimize patient contact with therapists, especially during March 2020 widespread lockdowns.

### Blinding and randomisation

The study was conducted under double-blind conditions, ensuring that investigators, study personnel, and patients remained unaware of treatment allocations. Given the amber color of Cerebrolysin, colored infusion lines were utilized during drug administration. Randomization was performed 1:1 in blocks of 4, and envelopes esignated for each enrolled patient were handed to the study nurses responsible for preparing the infusion solution, who were excluded from other study-related procedures and were strictly instructed not to disclose any information about treatment allocation. Treatment envelopes were only opened once the patient’s initial infusion was ready. Patients who met the inclusion and exclusion criteria were assigned a random number from a pre-generated list by a selected biometrician. Based on this list, sealed, opaque randomization/emergency envelopes were distributed (1) to the study center, for instances where unblinding was necessary due to potential harm to a patient; (2) to the individual preparing the infusion; (3) to the study coordinator. Upon opening, the date and time were recorded on the randomization/emergency envelopes, and they were signed by the individual who opened them. Any premature unblinding of the investigational products was immediately documented by the investigator and communicated to the coordinator. The entire study was unblinded after the database was closed and the analysis populations were determined.

### Evaluations

Western Aphasia Battery is an instrument for assessing the language function of adults with suspected neurological disorders because of a stroke, head injury, or dementia.^20^ It aids in identifying the presence, severity, and type of aphasia, while also establishing a baseline for monitoring changes during therapy, identifying key areas in the language capabilities of the patient to be improved during treatment, and inferring the location of the lesion that caused aphasia. The WAB targets English speaking adults and teens with a neurological disorder between the ages of 18 and 89 years old. The WAB tests both linguistic skills (such as speech, fluency, auditory comprehension, reading and writing), as well as non-linguistic skills (such as drawing, calculation, block design and apraxia). This study used a Romanian linguistically validated version of WAB.^21^

The National Institutes of Health Stroke Scale assesses neurologic deficit and is a 15 items scale that covers the level of consciousness, gaze, visual fields, facial palsy, motor functions, limb ataxia, aphasia, dysarthria and extinction and inattention.^22^ The NIHSS is observer-rated and takes 5-8 min to complete. Items have 3- to 5-point response scales, scored from 0 to 4 with higher score indicative of more severe disability. In case of patient death, the worst score possible will assigned. The NIHSS will be used to assess the severity of the stroke at baseline as well as in the follow-up examinations as a measure of neurological function deficit.

The Modified Rankin Score is a functional outcome scale measuring global outcome.^23^ It is used for grading the outcome and the level of disability after a stroke. The Modified Rankin Score is a 7-point ordinal scale with a score of 0 indicative of no residual symptoms at all and the worst possible score of 6 which is assigned in case of death. The Modified Rankin Scale is observer rated and takes about 5 min to complete.

The Barthel Scale/Index (BI) is an ordinal scale used to measure performance in activities of daily living (ADL).^24,25^ Ten variables describing ADL and mobility are scored, a higher number being a reflection of greater ability to function independently following hospital discharge. Time taken and physical assistance required to perform each item are used in determining the assigned value of each item. The Barthel Index measures the degree of assistance required by an individual on 10 items of mobility and self care ADL.

### Study endpoints

The primary objective of the ESCAS study was to assess the efficacy of Cerebrolysin and speech therapy versus placebo (saline solution) and speech therapy at 30, 60, and 90 days after baseline. Efficacy was evaluated using the WAB (Romanian translated version) scores at each time point. The primary outcome measure was the change in WAB score from baseline to each follow-up time point (30, 60, and 90 days).

The secondary objectives of the ESCAS study included: (1) To assess the efficacy of Cerebrolysin and speech therapy versus placebo (saline solution) and speech therapy at 30, 60, and 90 days after baseline using measures of motor, neurological, and global functional outcome. These outcomes were evaluated using the NIHSS, BI, and mRS scores at each time point; (2) To evaluate the safety of Cerebrolysin and speech therapy versus placebo (saline solution) and speech therapy at 30, 60, and 90 days after baseline. Safety was assessed by comparing the incidence of adverse events and severe adverse events between the two treatment groups. Additionally, we measured specific cardiovascular (such as, but not limited to stroke, myocardial infarction, atherosclerosis, vascular stenosis, as well as their recurrence), hematological (including anemia and vitamin B9 or B12 deficiency), renal system (including hyperuremia, hyperuricemia and urinary tract infections) and metabolic (including dyslipidemia, diabetes mellitus and atherosclerosis) related adverse events. The secondary outcome measures included changes in NIHSS, BI, and mRS scores from baseline to each follow-up time point (30, 60, and 90 days), as well as the incidence of AEs and SAEs.

### Study populations

The statistical analysis was done separately and was reported for each of the following study populations. Assessment of inclusion and exclusion was done independently for each patient and each analysis set. Each patient was included in all the relevant analysis sets. The Per Protocol population (PP) included all trial participants who adhered to the study protocol, received the medication corresponding to the group they were assigned to after randomization, had at most minimal protocol deviations, and no missing values for total scores of Western Aphasia Battery and NIHSS at visits 2, 3, or 4, or missing values for modified Rankin Scale and Barthel Index at visits 3 or 4. This analysis set was used to assess the efficacy of the treatment under ideal conditions. From a missing data handling perspective, the PP population was subject to listwise deletion, meaning that a patient missing one data point was not considered in any analysis. The Intention To Treat (ITT) population included all trial participants who were registered in the trial and were randomized, regardless of their subsequent adherence to the protocol or premature discontinuation. This analysis set was used to assess the efficacy of the studied treatment, taking into account patients not adhering to the protocol (non-compliance, dropouts, SAEs, unforeseen events), thus better representing the expected results of the treatment in clinical practice. From a missing data handling perspective, the ITT population was subject to pairwise deletion, meaning that a patient missing one data point was still considered in any analysis that did not require the missing data point. As such, compared to PP, ITT contained all the PP subjects, as well as subjects with major protocol deviations. The same statistical procedures were applied to both PP and ITT populations, with the difference lying in the selection of patients based on subsetting the database. The Safety Population (SP) included all trial participants who were registered in the trial and received at least one dose of treatment, regardless of their subsequent adherence to the protocol or premature discontinuation. Unlike ITT, SP analysis focused on AEs and SAEs caused by the treatment, thus evaluating the safety-related parameters of the products. Patients were included in this population irrespective of missing data or premature discontinuation.

### Sample size estimation

Preceding the clinical trial, we determined the sample size for the ESCAS study through a power analysis using G*Power 3.1.9.7^26^. Assuming a medium effect size d=0.5, an alpha error of α=0.05 and β=0.2, with an allocation ratio of 1 for Wilcoxon-Mann-Whitney test between two groups, we obtained a sample size of 53 patients per group. To provide a margin of error for situations such as patient withdrawal, SAEs or protocol violations, we have decided to recruit at least 120 patients in total for our study, 60 for each arm.

### Data analysis

We used Microsoft Excel 2019, (Microsoft Corporation, Redmond, WA), for data preparation and cleanup and R v. 4.3.1^27^ (R Core Team, Vienna, Austria) and RStudio^28^ (Posit Software, PBC, Boston, MA) for data analysis, loading the ggplot2^29^, readxl^30^, and xlsx^31^ libraries. For comparing numeric values of paired samples, differential values for each pair of values were computed. Then, the distribution of those differentials was compared with the normal distribution, using the Shapiro-Wilk test. If the differentials were normally distributed, we used a one-sample Student T test (equivalent with the Student T test for paired samples) with null hypothesis μ=0. If the differentials were not normally distributed, we used a Wilcoxon signed-rank test with null hypothesis location=0. For comparing numeric values of unpaired samples, the normality of the values in each sample was tested using the Shapiro-Wilk test. If values from both samples were normally distributed, then variance equality between the samples was assessed using the Bartlett test. If the variances were equal, then the differences between the two samples were assessed using a Student T test for unpaired samples and equal variances. If the variances were not equal, then a Student T test for unpaired samples and unequal variances was used. In both cases, the null hypothesis was that the mean difference was equal to 0. If values from at least one of the samples were not normally distributed, then a Wilcoxon rank sum with null hypothesis of location difference equal to 0 was performed. For comparing ordinal values of paired samples, a Wilcoxon signed-rank test was used, and for unpaired samples, a Wilcoxon rank-sum test was used. For comparing the difference in prevalences of variants of one dichotomous or nominal variable, among the groups of another dichotomous or nominal variable (i.e., testing the association between two dichotomous/nominal variables), the Chi^2^ test was used or, where its assumptions were violated (mainly due to a small number of patients in any group), the Fisher exact test was used. Where applicable, the two-tail p-value was reported. The type 1 error was assumed to be α=0.05, and as such, results were considered statistically significant for p<0.05. The full statistical analysis protocol was published before database unblinding.^19^

## Results

The study enrolled 132 patients, of which 123 meeting criteria for the ITT population (visit 2 Cerebrolysin n=61, placebo n=63; visit 3 Cerebrolysin n=59, placebo n=63; visit 4 Cerebrolysin n=58, placebo n=62), and 120 for the PP population (Cerebrolysin n=58, placebo n=62). A flow diagram of patient enrolment is available in Figure 1.

**Figure 1.**
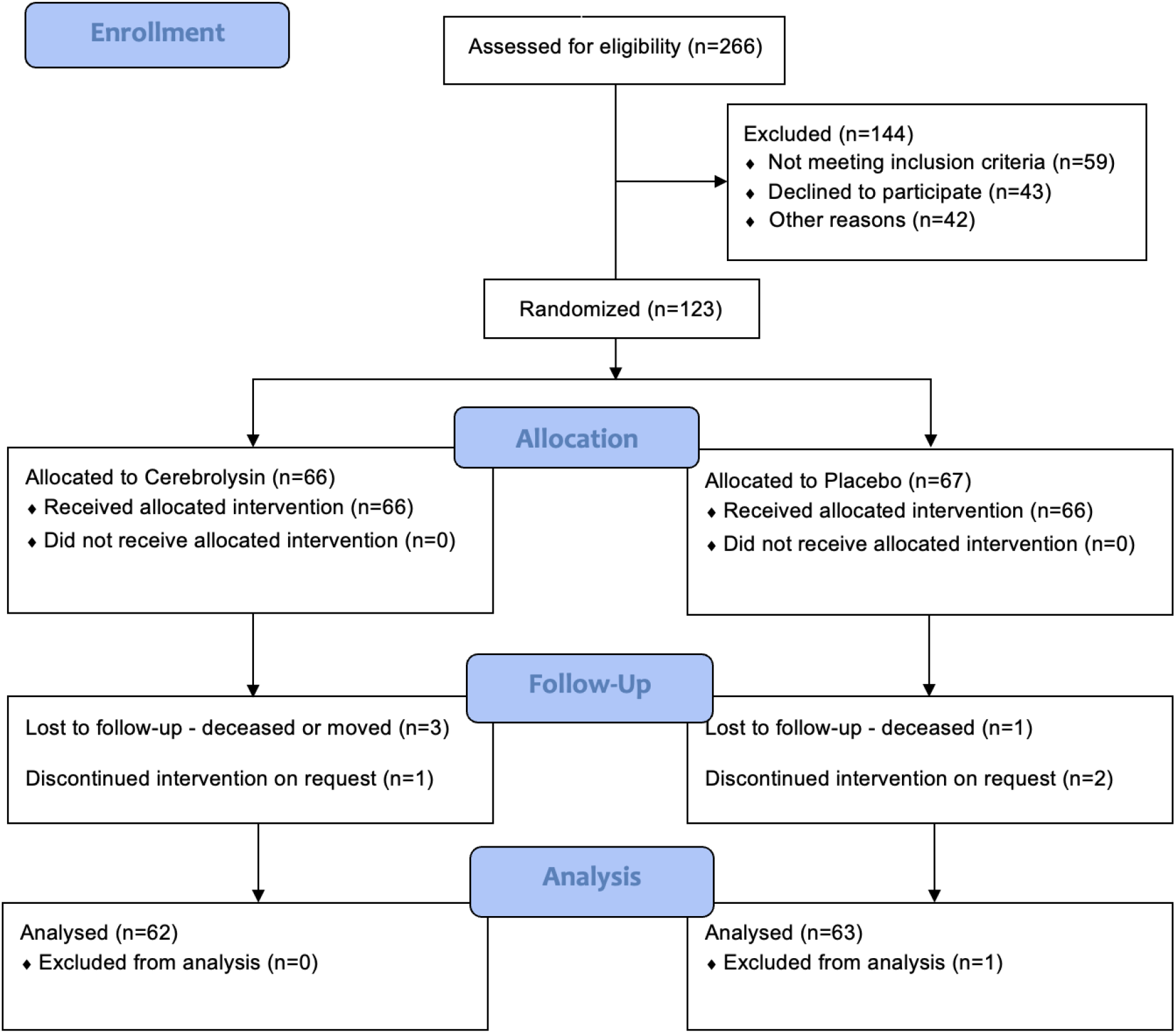
CONSORT flow diagram for the Per Protocol analysis

Demographic characteristics and baseline outcome measures highlighting good baseline comparability is available in Table 1.

**Table 1.** Demographic characteristics and baseline values for trial arms. SD standard deviation; Q1 quantile 1; Q3 quantile 3; † Wiloxon rank sum test; ⸸ Chi^2^ test; ‡ Fisher test.

| Variable | Indicator | Cerebrolysin | Placebo | p-value |
| --- | --- | --- | --- | --- |
|  | n = | 66 (50%) | 66 (50%) |  |
| Age | Mean±SD | 70.5±11.1 | 67.6±10.9 | 0.117† |
|  | Median (Q1 – Q3) | 73.5 (62.2 - 81) | 69.5 (62 - 76) |  |
| Gender | Female | 29 (43.9%) | 31 (46.9%) | 0.727‡ |
|  | Male | 37 (56.0%) | 35 (53.0%) |  |
| Education | No formal education | 0 (0%) | 0 (0%) | 0.351‡ |
|  | Primary school | 4 (6.0%) | 5 (7.5%) |  |
|  | Secondary school | 16 (24.2%) | 13 (19.6%) |  |
|  | High school | 40 (60.6%) | 35 (53.0%) |  |
|  | University | 6 (9.0%) | 13 (19.6%) |  |
| Alcohol use | None | 62 (93.9%) | 66 (100%) | 0.119‡ |
|  | Regular use | 4 (6.0%) | 0 (0%) |  |
|  | Abuse | 0 (0%) | 0 (0%) |  |
| Other substances use | None | 64 (96.9%) | 65 (98.4%) | 1‡ |
|  | Use | 2 (3.0%) | 1 (1.515%) |  |
|  | Abuse | 0 (0%) | 0 (0%) |  |
| WAB Aphasia quotient | Mean±SD | 50.7±19.8 | 56.021±22.442 | 0.154† |
|  | Median (Q1 – Q3) | 46.8 (34.8 - 64.2) | 53.1 (39.3 - 78.9) |  |
| NIHSS | Mean±SD | 9.2±4.4 | 8.5±4.3 | 0.362† |
|  | Median (Q1 – Q3) | 10 (6 - 12.7) | 8 (5 - 12) |  |

### ITT Efficacy Analysis - Western Aphasia Battery - Aphasia Quotient - baseline differences

For each patient, we analyzed the differences between WAB values at subsequent visits and WAB values at baseline. For the second visit, there were 61 valid data points for the Cerebrolysin group, with a median (quantile 1 – quantile 3) increase of 14.2 (10.3 – 20.5), statistically significant compared to baseline (p<0.001); in the placeo group, there were 63 valid data points, with a median increase of 8 (4.65 – 12.3), statistically significant compared to baseline (p<0.001). There was a higher increase in scores compared to baseline in the Cerebrolysin group than in the placebo group, which was statistically significant (p<0.001).

For the third visit, the Cerebrolysin group had 59 valid data points with a median increase of 27.3 (17 – 38.1) which was statistically significantly higher compared to baseline (p<0.001), and the placebo group had 63 valid data points with a median increase of 12.2 (8 - 22) which was statistically significantly higher compared to baseline (p<0.001). WAB score increases were statistically significantly higher in the Cerebrolysin group compared to placebo (p<0.001).

During the fourth visit, the cerebrolysin group had 58 valid data points with a median increase of 36.05 (20.8 - 47.35) which was statistically significantly higher compared to baseline (p<0.001), and the placebo group had 62 valid data points with a median increase of 17.1 (12.9 - 29.23) which was statistically significantly higher compared to baseline (p<0.001). WAB score increases were statistically significantly higher in the Cerebrolysin group compared to placebo (p<0.001).

More detailed information, including data expressed as mean and standard deviation and the statistical tests applied, is available in Supplementary File S1, Sheet 01.

### ITT Efficacy Analysis - NIHSS

Due to the fact that lower values of NIHSS are expected in relation to clinical recovery, we computed the differences between NIHSS scores at baseline and subsequent visits. During the second visit, the Cerebrolysin group had 61 valid data points with a median decrease of 3 (2 – 4) which was statistically significant compared to baseline values (p<0.001), while the placebo group had 63 valid data points with a median decrease of 2 (1-3) which was statistically significant compared to baseline (p<0.001). NIHSS scores had a statistically significantly higher decrease in the Cerebrolysin group (p=0.034).

For the third visit, the Cerebrolysin group had 59 valid data points with a median decrease of 5 (2.5 – 6.5), statistically significant compared to baseline (p<0.001), and the placebo group had 63 valid data points with a median decrease of 3 (2 – 4), statistically significant compared to baseline (p<0.001). There was a higher decrease in NIHSS scores compared to baseline in the Cerebrolysin group than in the placebo group, which was statistically significant (p<0.001).

During the fourth visit, the cerebrolysin group had 58 valid data points with a median decrease of 6 (3.25 – 8), statistically significant compared to the baseline (p<0.001), and the placebo group had 62 valid data points with a median decrease of 4 (2 – 5), statistically significant compared to the baseline (p<0.001). There were statistically significantly higher decreases in the Cerebrolysin group compared to the placebo group (p<0.001).

More detailed information, including data expressed as mean and standard deviation and the statistical tests applied, is available in Supplementary File S1, Sheet 02.

### ITT Efficacy Analysis - mRS

Modified Rankin Scale absolute values at second, third and fourth visits were reported. During the second visit, the Cerebrolysin group consisted of 61 valid data points with a median of 3 (2 – 4), while the placebo group had 63 valid data points with a median of 3 (2 – 4). There were no statistically significant differences in mRS values between the group treated with Cerebrolysin and placebo at visit 2 (p=0.492).

For the third visit, the cerebrolysin group had 59 valid data points with a median of 2 (1 – 3), and the placebo group had 63 valid data points with a median of 3 (2 – 4). Both groups showed statistically significant decreases in mRS values compared to the corresponding values from visit 2 (p<0.001 for both groups). Moreover, the decreases compared to the second visit were statistically significantly higher in the Cerebrolysin group compared to placebo (p=0.005).

During the fourth visit, the Cerebrolysin group had 58 valid data points with a median of 2 (1 – 2), and the placebo group had 62 valid data points with a median of 2 (1 – 3). Both groups showed statistically significant decreases in mRS values compared to the corresponding values from visit 2 (p<0.001 for both groups). Moreover, the decreases compared to the second visit were statistically significantly higher in the Cerebrolysin group compared to placebo (p<0.001).

More detailed information, including data expressed as mean and standard deviation and the statistical tests applied, is available in Supplementary File S1, Sheet 03.

### ITT Efficacy Analysis - BI

Barthel Index absolute values at second, third and fourth visits were reported. During the second visit, the Cerebrolysin group consisted of 61 valid data points with a median of 60 (40 - 90), while the placebo group had 63 valid data points with a median of 65 (45 - 95). There were no statistically significant differences in BI values between the group treated with Cerebrolysin and placebo at visit 2 (p=0.81).

For the third visit, the Cerebrolysin group had 59 valid data points with a median of 70 (55 - 100), and the placebo group had 63 valid data points with a median of 70 (55 - 92.5). Both groups showed statistically significant increases in BI values compared to the corresponding values from visit 2 (p<0.001 for both groups). Moreover, the increases compared to the second visit were statistically significantly higher in the Cerebrolysin group compared to placebo (p=0.008).

During the fourth visit, the Cerebrolysin group had 58 valid data points with a median of 85 (75 - 100), and the placebo group had 62 valid data points with a median of 80 (56.25 - 95). Both groups showed statistically significant increases in mRS values compared to the corresponding values from visit 2 (p<0.001 for both groups). Moreover, the increases compared to the second visit were statistically significantly higher in the Cerebrolysin group compared to placebo (p<0.001).

More detailed information, including data expressed as mean and standard deviation and the statistical tests applied, is available in Supplementary File S1, Sheet 04.

### PP Efficacy Analysis - Western Aphasia Battery - Aphasia Quotient - baseline differences (Figure 2)

In the PP dataset, there were 58 valid data points in the Cerebrolysin group and 62 valid data points in the placebo group, available for all visits and all measurements.

**Figure 2.**
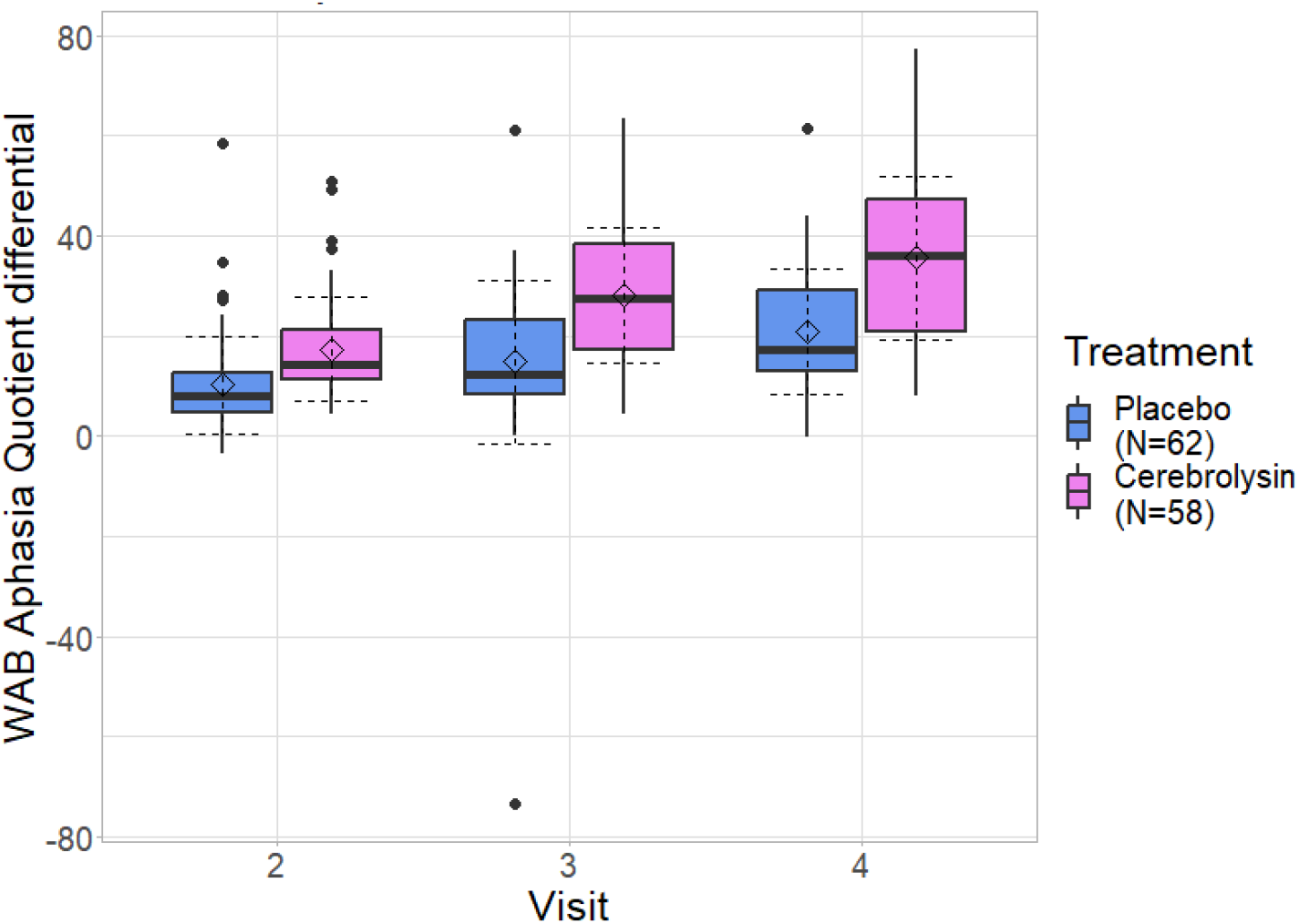
Boxplot representing distribution of Western Aphasia Battery (WAB) – Aphasia Quotient differences compared to baseline (Per Protocol population); boxes indicate interquartile range, whiskers represent range, horizontal lines represent medians, dashed diamonds represent mean and dashed error lines represent the standard deviation relative to the mean.

During the second visit, the Cerebrolysin group had a median increase in WAB scores of 14.35 (11.4 - 21.03) which was statistically significant compared to baseline (p<0.001), while the placebo group had a median increase of 8 (4.75 - 12.75) which was also statistically significant compared to baseline (p<0.001). The increase in WAB scores was significantly higher in the Cerebrolysin group, compared to placebo (p<0.001).

During the third visit, the Cerebrolysin group had a median increase in WAB scores of 27.35 (17.25 - 38.45) which was statistically significant compared to baseline (p<0.001), while the placebo group had a median increase of 12.4 (8.325 - 23.1) which was also statistically significant compared to baseline (p<0.001). The increase in WAB scores was significantly higher in the Cerebrolysin group, compared to placebo (p<0.001).

During the fourth visit, both the CRB and PLC groups had 58 and 62 valid data points, respectively. The mean score for the CRB group was 35.6 (SD = 16.3), and for the PLC group, it was 20.8 (SD = 12.4). The median score for the CRB group was 36.1, with values ranging from 20.8 to 47.3, and for the PLC group, it was 17.1, ranging from 12.9 to 29.2. The Cerebrolysin group showed significant differentials with p-values less than 0.001 as compared to PLC.

More detailed information, including data expressed as mean and standard deviation and the statistical tests applied, is available in Supplementary File S1, Sheet 05.

### PP Efficacy Analysis - NIHSS - baseline differences (Figure 3)

We computed the differences between NIHSS scores at baseline and subsequent visits. During the second visit, the Cerebrolysin group had a median decrease of 3 (2 – 4) which was statistically significant compared to baseline values (p<0.001), while the placebo group had a median decrease of 2 (1-3) which was statistically significant compared to baseline (p<0.001). NIHSS scores had a statistically significantly higher decrease in the Cerebrolysin group compared to placebo (p=0.019). For the third visit, the Cerebrolysin group had a median decrease of 5 (3 – 6.8), statistically significant compared to baseline (p<0.001), and the placebo group had a median decrease of 3 (2 – 4), statistically significant compared to baseline (p<0.001). NIHSS scores had a statistically significantly higher decrease in the Cerebrolysin group compared to placebo (p<0.001).

**Figure 3.**
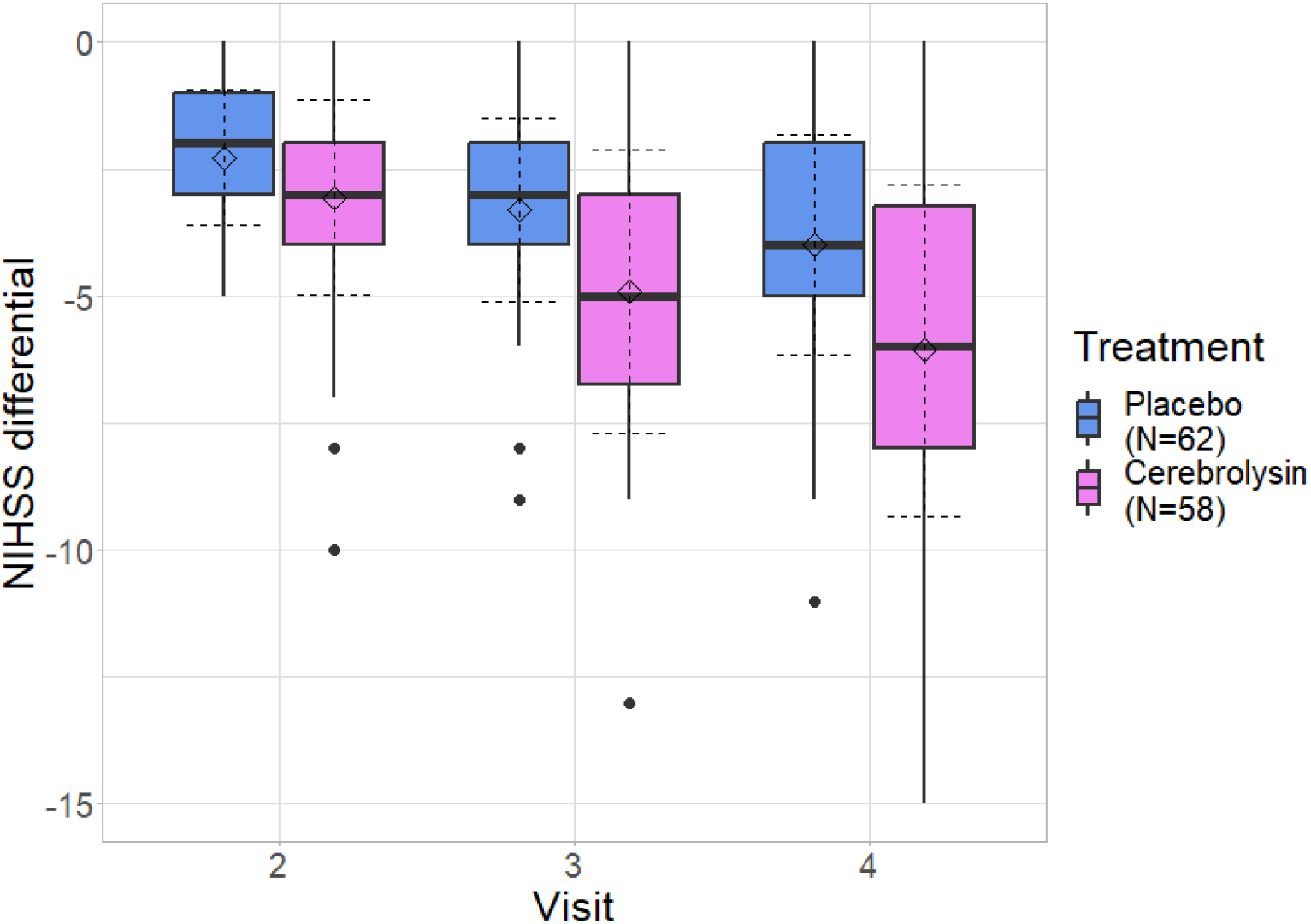
Boxplot representing distribution of National Institutes of Health Stroke Scale (NIHSS) score differences compared to baseline (Per Protocol population); boxes indicate interquartile range, whiskers represent range, horizontal lines represent medians, dashed diamonds represent mean and dashed error lines represent the standard deviation relative to the mean.

During the fourth visit, the cerebrolysin group had a median decrease of 6 (3.25 – 8), statistically significant compared to the baseline (p<0.001), and the placebo group had a median decrease of 4 (2 – 5), statistically significant compared to the baseline (p<0.001). There were statistically significantly higher decreases in the Cerebrolysin group compared to the placebo group (p<0.001).

More detailed information, including data expressed as mean and standard deviation and the statistical tests applied, is available in Supplementary File S1, Sheet 06.

### PP Efficacy Analysis - mRS - baseline differences (Figure 4)

During the second visit, the Cerebrolysin group had median mRS values of 3 (2 – 4), while the placebo group had a median of 3 (2 – 4). There were no statistically significant differences in mRS values between the group treated with Cerebrolysin and placebo at visit 2 (p=0.673).

**Figure 4.**
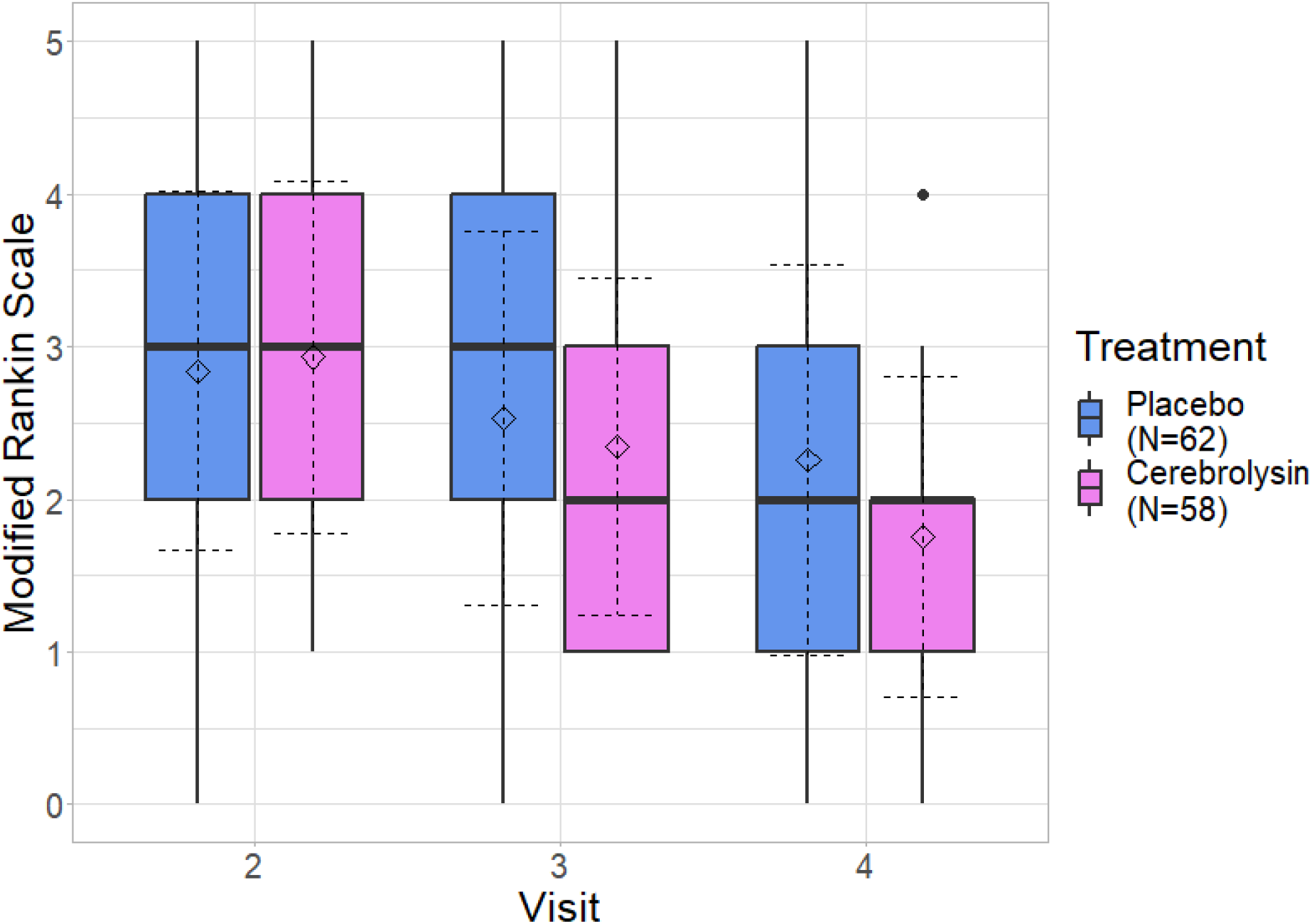
Boxplot representing distribution of modified Rankin Scale scores (Per Protocol population); boxes indicate interquartile range, whiskers represent range, horizontal lines represent medians, dashed diamonds represent mean and dashed error lines represent the standard deviation relative to the mean.

For the third visit, the cerebrolysin group had a median mRS of 2 (1 – 3), and the placebo group had a median of 3 (2 – 4). Both groups showed statistically significant decreases in mRS values compared to the corresponding values from visit 2 (p<0.001 for both groups). Moreover, the decreases were statistically significantly higher in the Cerebrolysin group compared to placebo (p=0.005).

During the fourth visit, the Cerebrolysin group had a median mRS of 2 (1 – 2), and the placebo group had a median of 2 (1 – 3). Both groups showed statistically significant decreases in mRS values compared to the corresponding values from visit 2 (p<0.001 for both groups). Moreover, the decreases compared to the second visit were statistically significantly higher in the Cerebrolysin group compared to placebo (p<0.001).

More detailed information, including data expressed as mean and standard deviation and the statistical tests applied, is available in Supplementary File S1, Sheet 07.

### PP Efficacy Analysis - BI - baseline differences (Figure 5)

During the second visit, the Cerebrolysin group had BI values with a median of 60 (40 - 90), while the placebo group had a median of 65 (45 - 95). There were no statistically significant differences in BI values between the group treated with Cerebrolysin and placebo at visit 2 (p=0.883).

**Figure 5.**
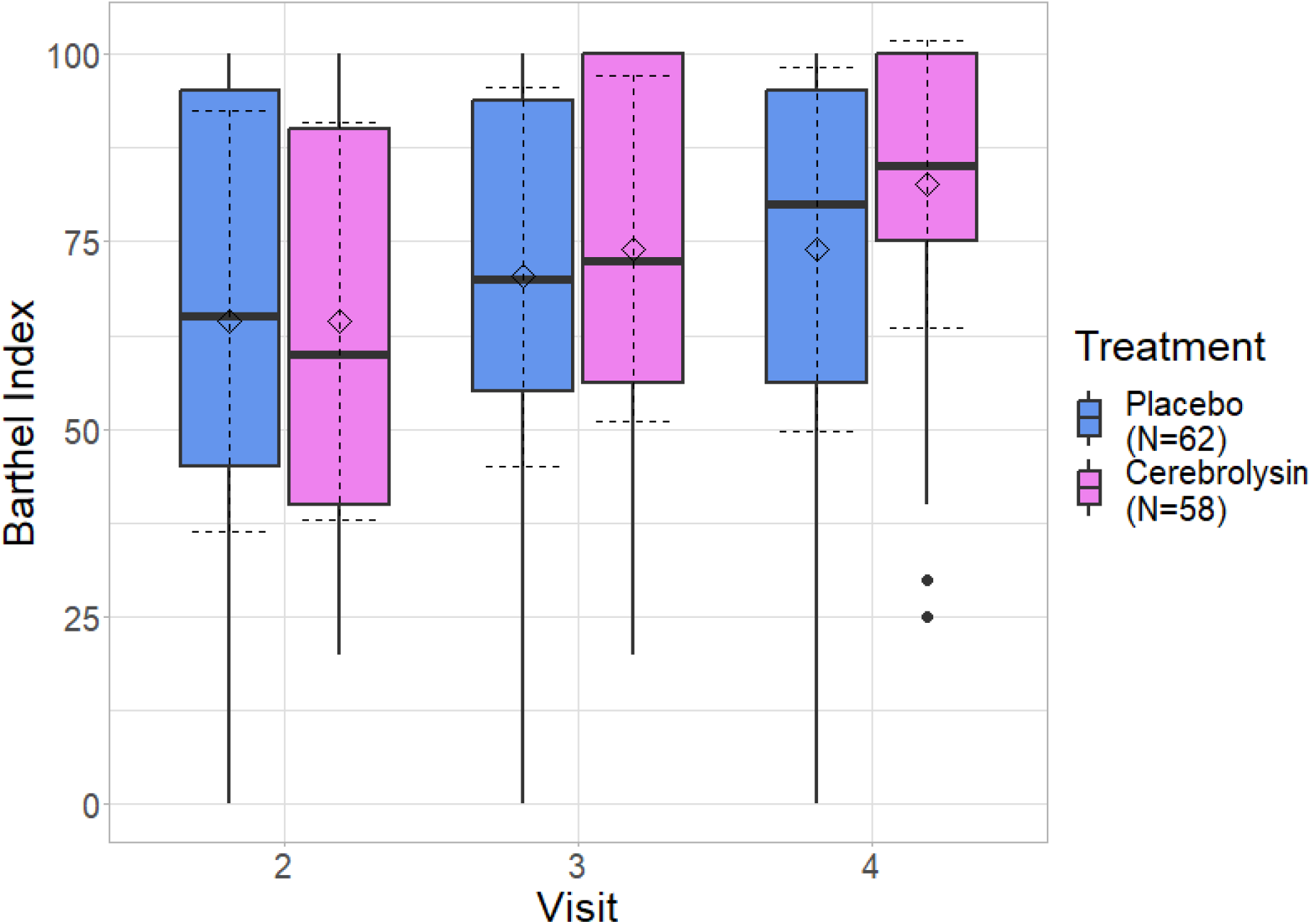
Boxplot representing distribution of Barthel Index scores (Per Protocol population); boxes indicate interquartile range, whiskers represent range, horizontal lines represent medians, dashed diamonds represent mean and dashed error lines represent the standard deviation relative to the mean. BI baseline differences (PP); boxes indicated interquartile range, whiskers represent range, horizontal lines represent averages.

For the third visit, the Cerebrolysin group had a median of 72.5 (56.25 - 100), and the placebo group had a median of 70 (55 - 93.75). Both groups showed statistically significant increases in BI values compared to the corresponding values from visit 2 (p<0.001 for both groups). Moreover, the increases were statistically significantly higher in the Cerebrolysin group compared to placebo (p=0.007).

During the fourth visit, the Cerebrolysin group had a median of 85 (75 - 100), and the placebo group had a median of 80 (56.25 - 95). Both groups showed statistically significant increases in mRS values compared to the corresponding values from visit 2 (p<0.001 for both groups). Moreover, the increases compared to the second visit were statistically significantly higher in the Cerebrolysin group compared to placebo (p=0.002).

More detailed information, including data expressed as mean and standard deviation and the statistical tests applied, is available in Supplementary File S1, Sheet 08.

### Safety analysis

Regarding adverse events, there was no significant difference in the number of patients experiencing SAEs between the two groups (p=0.381). For general AEs, 43.9% of patients in the Cerebrolysin group and 30.3% in the placebo group reported events, although the difference was not statistically significant (p=0.105). The mean number of AEs per patient was 0.894 in the Cerebrolysin group and 0.636 in the placebo group (no difference between the number of AEs experienced by each patient between groups, p=0.134). Specifically, for AE categories such as neurological, psychiatric, respiratory, immune-related, and ophthalmic, both groups showed similar rates with no significant differences. Highest differences in AEs reported between groups were seen in the gastrointestinal category (4.6% of the Cerebrolysin group reported AEs, while none were reported in the placebo group - p=0.244) and in the osteoarticular category (no AEs were reported in the Cerebrolysin group, while 6.1% of the patients in the placebo group reported events - p=0.119). Additional safety information is available in the Supplementary File S1, Sheet 09.

## Discussion

The ESCAS study sought to evaluate the efficacy and safety of Cerebrolysin in combination with speech therapy compared to a placebo (saline solution) in combination with speech therapy for the treatment of aphasia following acute ischemic stroke. Aphasia, a debilitating consequence of stroke, significantly impacts an individual’s quality of life and ability to communicate effectively. While SLT has long been the gold standard for aphasia rehabilitation, this study aimed to explore the potential synergistic effects of Cerebrolysin as an adjunctive therapy to enhance recovery in patients with persistent aphasia.

The demographic characteristics and baseline outcome measures revealed good baseline comparability between the Cerebrolysin and placebo groups, ensuring that any observed differences in outcomes could be attributed to the interventions rather than baseline disparities.

The primary outcome measure in this study was the change in WAB scores from baseline to each follow-up time point at 30, 60, and 90 days. The results demonstrated significant improvements in the WAB Aphasia Quotient for both the Cerebrolysin and placebo groups at each time point compared to baseline. These findings indicate that speech therapy alone, as part of the standard care, led to notable improvements in aphasia over time. However, it is important to note that the improvements observed in the Cerebrolysin group were consistently significantly higher than those in the placebo group, with large numeric differences. This provides direct evidence supporting the addition of Cerebrolysin to SLT in the post-stroke aphasia neurorehabilitation regimen.

Secondary outcome measures, including the NIHSS, BI, mRS, were also assessed to evaluate motor, neurological, and global functional outcomes. Similar to the WAB scores, the placebo group showed significant improvements in these measures at each time point compared to baseline, indicating that speech therapy alone had a positive impact on overall recovery. Still, significantly higher improvements were seen across all measurements in the Cerebrolysin group, compared to those in the placebo group, thus further supporting the role of Cerebrolysin in the functional recovery of the post-stroke patient.

The safety analysis included the assessment of adverse events and severe adverse events in both treatment groups. No specific safety concerns related to Cerebrolysin were identified during the study. The incidence of AEs and SAEs was similar between the Cerebrolysin and placebo groups, suggesting that the addition of Cerebrolysin to speech therapy did not result in an increased risk of adverse events, adding to the existing body of evidence available on the safety of Cerebrolysin^14,17,18,32^

The results of this study provide valuable insights into the potential benefits of adding Cerebrolysin to speech therapy for the rehabilitation of post-stroke aphasia. The trends observed suggest that Cerebrolysin may have a positive impact on aphasia rehabilitation. These findings warrant further investigation in larger studies with a longer follow-up period to determine whether Cerebrolysin can produce statistically significant improvements in aphasia outcomes.

It is essential to acknowledge some limitations of this study. The sample size, although determined through a rigorous power analysis, may have limited the ability to detect small but clinically still meaningful differences between the treatment groups.

In conclusion, the ESCAS trial represents an important step in exploring Cerebrolysisn as an adjunctive therapy, for aphasia rehabilitation following acute ischemic stroke. The trends toward improved outcomes with Cerebrolysin suggest the need larger clinical trials to confirm our result. Aphasia remains a challenging condition, and finding additional therapies that can enhance recovery and quality of life for affected individuals is of paramount importance. Future studies with larger sample sizes and longer follow-up durations may provide more definitive answers regarding the efficacy of Cerebrolysin in aphasia rehabilitation.

## Data Availability

Data on which analyses have been performed is available as supplementary material. Further data requests may be made to the corresponding authors.

## Acknowledgements

The authors would like to acknowledge the contributions of team members involved in investigation of this project: Dr. Hanna Dragos, Dr. Flavius Dan, Dr. Georgiana Munteanu, Psych. Bianca Bora, Ms. Diana Chira.

## References

1. Huppertz V, Guida S, Holdoway A, Strilciuc S, Baijens L, Schols JMGA, van Helvoort A, Lansink M, Muresanu DF. Impaired Nutritional Condition After Stroke From the Hyperacute to the Chronic Phase: A Systematic Review and Meta-Analysis. Front Neurol. 2021;12:780080.

2. Feigin VL, Stark BA, Johnson CO, Roth GA, Bisignano C, Abady GG, Abbasifard M, Abbasi-Kangevari M, Abd-Allah F, Abedi V, et al. Global, regional, and national burden of stroke and its risk factors, 1990–2019: a systematic analysis for the Global Burden of Disease Study 2019. The Lancet Neurology. 2021;20:795–820.

3. Brady MC, Kelly H, Godwin J, Enderby P, Campbell P. Speech and language therapy for aphasia following stroke. Cochrane Database Syst Rev. 2016;CD000425.

4. Berthier ML. Poststroke aphasia : epidemiology, pathophysiology and treatment. Drugs Aging. 2005;22:163–182.

5. Butler RA, Lambon Ralph MA, Woollams AM. Capturing multidimensionality in stroke aphasia: mapping principal behavioural components to neural structures. Brain. 2014;137:3248–3266.

6. Brady MC, Ali M, VandenBerg K, Williams LJ, Williams LR, Abo M, Becker F, Bowen A, Brandenburg C, Breitenstein C, et al. Complex speech-language therapy interventions for stroke-related aphasia: the RELEASE study incorporating a systematic review and individual participant data network meta-analysis [Internet]. Southampton (UK): National Institute for Health and Care Research; 2022 [cited 2024 Feb 19]. Available from: http://www.ncbi.nlm.nih.gov/books/NBK584830/

7. Marebwa BK, Fridriksson J, Yourganov G, Feenaughty L, Rorden C, Bonilha L. Chronic post-stroke aphasia severity is determined by fragmentation of residual white matter networks. Sci Rep. 2017;7:8188.

8. Muresanu DF. Neuroplasticity and Neurorecovery. Stroke. 2009;37–49.

9. Muresanu DF, Buzoianu A, Florian SI, von Wild T, Muresanu D. Towards a roadmap in brain protection and recovery. J Cell Mol Med. 2012;16:2861–2871.

10. Muresanu DF, Heiss W-D, Hoemberg V, Bajenaru O, Popescu CD, Vester JC, Rahlfs VW, Doppler E, Meier D, Moessler H, et al. Cerebrolysin and Recovery After Stroke (CARS): A Randomized, Placebo-Controlled, Double-Blind, Multicenter Trial. Stroke. 2016;47:151–159.

11. Guekht A, Vester J, Heiss W-D, Gusev E, Hoemberg V, Rahlfs VW, Bajenaru O, Popescu BO, Doppler E, Winter S, et al. Safety and efficacy of Cerebrolysin in motor function recovery after stroke: a meta-analysis of the CARS trials. Neurol Sci. 2017;38:1761–1769.

12. Amiri-Nikpour MR, Nazarbaghi S, Ahmadi-Salmasi B, Mokari T, Tahamtan U, Rezaei Y. Cerebrolysin effects on neurological outcomes and cerebral blood flow in acute ischemic stroke. Neuropsychiatr Dis Treat. 2014;10:2299–2306.

13. Brainin M. Cerebrolysin: a multi-target drug for recovery after stroke. Expert Rev Neurother. 2018;18:681–687.

14. Mure?anu DF, Livin? Popa L, Chira D, Dăbală V, Hapca E, Vlad I, Văcăra? V, Popescu BO, Chereche? R, Strilciuc ?tefan, et al. Role and Impact of Cerebrolysin for Ischemic Stroke Care. Journal of Clinical Medicine [Internet]. 2022;11. Available from: https://www.mdpi.com/2077-0383/11/5/1273

15. Muresanu DF, Vester JC. Prospective Meta-Analysis (PMA) of the Cerebrolysin Trials in Neuroprotection and Neurorecovery after Traumatic Brain Injury (CAPTAIN I and CAPTAIN II) [Internet]. 2016;Available from: https://ssnn.ro/images/Images/Research/PMA15062016.pdf

16. Beghi E, Binder H, Birle C, Bornstein N, Diserens K, Groppa S, Homberg V, Lisnic V, Pugliatti M, Randall G, et al. European Academy of Neurology and European Federation of Neurorehabilitation Societies guideline on pharmacological support in early motor rehabilitation after acute ischaemic stroke. European Journal of Neurology. 28:2831–2845.

17. Strilciuc S. Safety of Cerebrolysin for neurorecovery after acute ischemic stroke: a systematic review and meta-analysis of twelve randomized-controlled trials. 2021 [cited 2021 Dec 3];Available from: https://osf.io/cxufq

18. Bornstein NM, Guekht A, Vester J, Heiss W-D, Gusev E, Hömberg V, Rahlfs VW, Bajenaru O, Popescu BO, Muresanu D. Safety and efficacy of Cerebrolysin in early post-stroke recovery: a meta-analysis of nine randomized clinical trials. Neurol. Sci. 2018;39:629–640.

19. Muresanu DF, Verisezan Rosu O. Can Cerebrolysin improve aphasia after ischemic stroke? [Internet]. [cited 2024 Feb 19];Available from: http://www.isrctn.com/ISRCTN54581790

20. Shewan CM, Kertesz A. Reliability and Validity Characteristics of the Western Aphasia Battery (WAB). J Speech Hear Disord. 1980;45:308–324.

21. Kory Calomfirescu S. Romanian version of Western Aphasia Battery. Afazia în accidentele vasculare cerebrale. 1996;74–95.

22. Meyer BC, Hemmen TM, Jackson CM, Lyden PD. Modified National Institutes of Health Stroke Scale for use in stroke clinical trials: prospective reliability and validity. Stroke. 2002;33:1261– 1266.

23. Van Swieten JC, Koudstaal PJ, Visser MC, Schouten HJ, Van Gijn J. Interobserver agreement for the assessment of handicap in stroke patients. Stroke. 1988;19:604–607.

24. Collin C, Wade DT, Davies S, Horne V. The Barthel ADL Index: a reliability study. Int Disabil Stud. 1988;10:61–63.

25. Mahoney FI, Barthel DW. FUNCTIONAL EVALUATION: THE BARTHEL INDEX. Md State Med J. 1965;14:61–65.

26. Faul F, Erdfelder E, Lang A-G, Buchner A. G*Power 3: a flexible statistical power analysis program for the social, behavioral, and biomedical sciences. Behav Res Methods. 2007;39:175– 191.

27. R Core Team. R: A Language and Environment for Statistical Computing [Internet]. Vienna, Austria: R Foundation for Statistical Computing; 2023. Available from: https://www.R-project.org/

28. Posit team. RStudio: Integrated Development Environment for R [Internet]. Boston, MA: Posit Software, PBC; 2023. Available from: http://www.posit.co/

29. Wickham H. ggplot2: Elegant Graphics for Data Analysis [Internet]. Springer-Verlag New York; 2016. Available from: https://ggplot2.tidyverse.org

30. Wickham H, Bryan J. readxl: Read Excel Files [Internet]. 2023. Available from: https://CRAN.R-project.org/package=readxl

31. Dragulescu A, Arendt C. xlsx: Read, Write, Format Excel 2007 and Excel 97/2000/XP/2003 Files [Internet]. 2020. Available from: https://CRAN.R-project.org/package=xlsx

32. Ziganshina LE, Abakumova T, Vernay L. Cerebrolysin for acute ischaemic stroke. Cochrane Database Syst Rev. 2016;12:CD007026.

